# Leveraging neighborhood-level Information to Improve Model Fairness in Predicting Prenatal Depression

**DOI:** 10.1101/2025.05.12.25327329

**Authors:** Yongchao Huang, Suzanne Alvernaz, Sage J. Kim, Pauline M. Maki, Andrew D. Boyd, Yang Dai, Beatriz Peñalver Bernabé

**Author notes:** co-senior authors Contact (Yang Dai, Beatriz Peñalver Bernabé).

## Abstract

**Importance:** Perinatal depression (PND) affects 10-20% of pregnant women, with significant racial disparities in prevalence, screening, and treatment. Neighborhood-level factors significantly influence PND risk, particularly among women of color, but current machine learning models using electronic medical records (EMRs) rarely incorporate neighborhood characteristics.

**Objective:** To determine whether integrating neighborhood-level information with EMRs improves fairness in PND prediction while identifying key neighborhood factors influencing model bias across racial/ethnic groups.

**Design, Setting, and Participants:** Study of 6,137 pregnant women who received care at a large urban academic hospital from 2010-2019, comprising 58% Non-Hispanic Black (NHB), 10% Non-Hispanic White (NHW), and 28% Hispanic (H) individuals, with depression status determined by PHQ-9 scores.

**Exposures:** 125 neighborhood-level factors from Chicago Health Atlas merged with 61 EMR features based on residential location.

**Main Outcomes and Measures:** Model performance (ROCAUC, PRAUC) and fairness metrics (disparate impact, equal opportunity difference, equalized odds). Feature importance analyzed using Shapley values and the impact of each neighborhood factor on model bias were evaluated. **Results** Models integrating neighborhood-level measures showed moderate predictive performance (ROCAUC: NHB 55%, NHW 57%, H 58%) while significantly improving fairness metrics compared to EMR-only models (p<0.05). Factors, such as suicide mortality rate and neighborhood safety rate, helped reduce bias. NHB women showed stronger correlations between PND risk factors and neighborhood variables compared to other groups. Most neighborhood factors had differential impacts across racial/ethnic groups, increasing bias for NHB women while reducing it for Hispanic women.

**Conclusions and Relevance:** Incorporating neighborhood-level information enhances fairness in PND prediction while maintaining predictive capability. The differential impact of neighborhood factors across racial/ethnic groups highlights the importance of considering neighborhood context in clinical risk assessment to reduce disparities in prenatal depression care.

## Introduction

Perinatal depression (PND), depression during pregnancy and up to one year postpartum, typically affects 10-20% of women annually^1^. PND can lead to severe consequences, including increased risk of pregnancy complications, preterm labor^2, 3^, maternal morbidity and mortality^4^, and adverse child developmental outcomes^4, 5^. Know risk factors for PND are: xxx. PND risk is also influenced by social determinants of health (SDoH), encompassing individual-level factors (such as racial/ethnic background and age) and neighborhood-level factors (such as green spaces, contamination, food access, and exposure to violence).^6-8^. These neighborhood-level factors may directly or indirectly affect mental health outcomes among vulnerable pregnant individuals via limited access to healthcare services and reduced social support networks^9^. Moreover, the interaction effect of individual- and neighborhood-level factors can amplify the risk of PND, potentially exacerbating health disparities among different racial and ethnic groups.

Social Neighborhood factors significantly influence PND risk^6, 10^, with individuals living in the most disadvantaged neighborhoods showing a 14% higher risk of PND compared to those in the least disadvantaged areas^11^. These neighborhood factors shape individual risk profiles and provide a more comprehensive understanding of PND risk across populations that live in different communities^7^. Non-Hispanic Black (NHB) women are disproportionately affected by the negative health impacts of SDoH and face a higher risk of PND compared to Non-Hispanic White (NHW) women ^6, 12, 13^, with the risk of PND increases in a dose-response manner from 39% to 60% as neighborhood disadvantage increases^11^. This disparity is closely linked to socioeconomic disadvantages, including poverty, housing instability, and correlates with adverse birth outcomes such as preterm delivery, low birth weight, and impaired fetal growth^14, 15^. The disparity is further exacerbated by lower rates of screening, treatment initiation, and follow-up care among women of color throughout the perinatal period^16^.

Current electronic medical record (EMR)-based machine learning (ML) models to predict PND rarely include neighborhood-level information^17^, which negatively affects women living in disadvantaged areas, where neighborhood factors (e.g., crime rate, pollution, long distances to groceries and hospitals) may play a crucial role in PND risk^18^. By focusing solely on individual-level factors (e.g., medical history, income, insurance status), EMR-based ML models may produce biased predictions and underestimate the risk for women exposed to multiple layers of social and environmental stressors ^19^. Addressing these disparities in developing fair ML models requires incorporating neighborhood-level data, as the effects of neighborhood disadvantage on PND risk vary widely across racial and ethnic groups^11, 20^.

In this study, we aim to enhance the assessment of early pregnancy depression risk by integrating social neighborhood information with EMRs. We employed EMRs from 6,137 pregnant women and combined them with a total of 125 neighborhood-level characteristics to determine the predictive and bias-reduction capabilities of the addition of neighborhood information. We seek to develop a more equitable model for evaluating depression symptom severity in early pregnancy.

## Materials and Method

### Study Population

EMRs from a total of 6,137 pregnant individuals were obtained from the University of Illinois Chicago (UIC) Hospital Health Sciences System, UIC Medical Center (UI Health). The population served at UI Health consists of 58% NHB, 10% NHW, 28% Hispanic, and 4% Asian and Native American. We extracted EMRs, including 694 features, from patients who were older than 17 years and who received obstetric services and gave birth at UI Health from 2010 to 2019. Depressive symptom severity was assessed using a standard self-reported patient health questionnaire-9^21^ at their first visit. The EMR extraction was approved by the University of Illinois Chicago, Institutional Review Board (IRB # 2020-0553).

### Preprocessing of EMRs and neighborhood information

EMRs were processed as previously described ^22^, including patients with PHQ-9 scores of 1-4 (controls) or ≥9 (cases) who were in early pregnancy (<24 weeks gestation), while excluding mild depression cases (scores 5-8) and those reporting no symptoms to minimize false negatives. (**Table S1**). Neighborhood level information expanding from 2010 to 2022 was obtained from a publicly available database, Chicago Health Atlas^23^, which contains more than 200 features. We included CHA data through 2022 to capture the complete temporal trends of neighborhood characteristics, as these features typically show stable patterns over time and provide a more comprehensive understanding of social neighborhood factors. Neighborhood information included data pertaining to morbidity, mortality, physical environment, clinical care, socioeconomic factors, health, and demography, just to name a few Addresses of each patient was mapped to the corresponding neighborhoods in Chicago rending a total of 77 neighborhood areas in Chicago and 260 neighborhood features (**Fig. 1**). The CHA data were averaged across time between 2010 to 2022. We removed neighborhood features that were missing in more than 5% of the neighborhoods to ensure data reliability. Additionally, when both raw counts and standardized rates were available for the same neighborhood feature, we used only the rate data and excluded the count data to avoid redundancy and better account for population differences across neighborhoods. After preprocessing, the dataset was composed of 61 EMR and 125 social neighborhood features for a total of 3,313 patients. Missing values from EMR and neighborhood information were imputed using the *MICE* package in Python (version 3.15.0)^24^. We sampled a total of 50 imputed datasets with 10 sampling iterations per imputed set to ensure the robustness of the imputed value. We merged 50 imputed datasets into one final dataset for analysis; details can be found in our prior study^22^.

**Figure 1.**
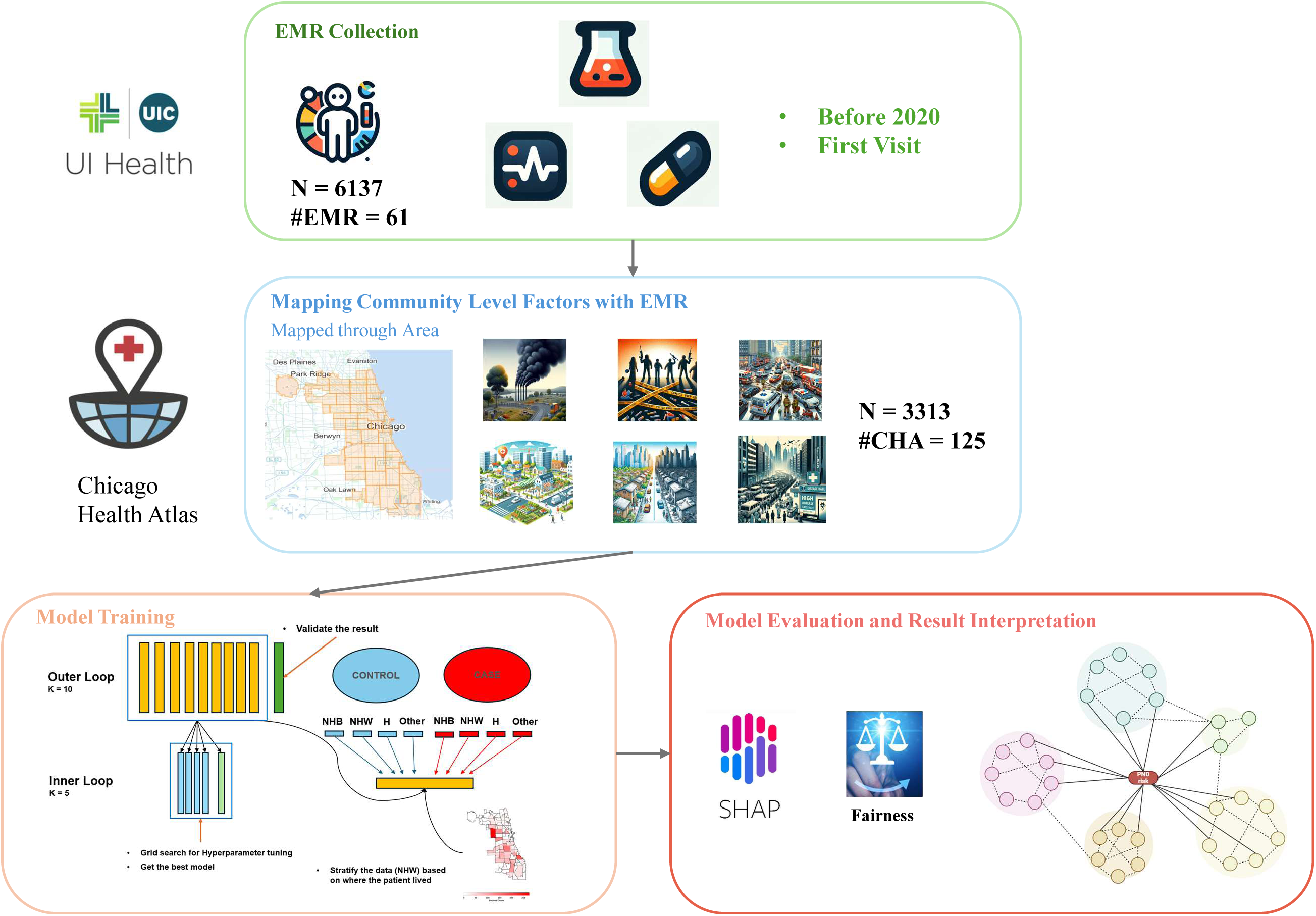
Study workflow for integrating EMR data with neighborhood-level factors for PND prediction. The workflow consists of three main components: a) EMR Collection: Data collection from EMR system (N=6,137) including patient data before 2020 and first visit information; b) Mapping Neighborhood Level Factors: Integration of neighborhood-level features with EMR records based on neighborhood areas, resulting in 3,313 patients and 125 neighborhood-level features; c) Model Development and Analysis: Including model training with nested cross-validation, stratified sampling by neighborhood, and evaluation using SHAP analysis and fairness metrics.

### Machine learning model selection and training

We evaluated three different machine learning models (Elastic Net^25^, Random Forest^26^, and XGBoost^27^) to predict PND. We selected the best model using nested k-fold cross-validation, optimizing the area under the receiver operating characteristic curve (ROCAUC) while maintaining a fixed sensitivity of 85% to maximize the true depression cases. Additional metrics used to evaluate model performance included: the area under the precision-recall curve (PRAUC), F1 score (harmonic mean of precision and recall), positive predictive value (PPV), negative predictive value (NPV), accuracy, and the standard deviation of true positive rate (TPR) and false positive rate (FPR). The nested k-fold cross-validation consisted of two parts: an outer loop that divided the data into training and test sets (k=10), and an inner loop (k=5) that further split the outer loop’s training set into multiple training and validation sets for model training and hyperparameter optimization (**Fig. S1**). To ensure representative sampling across subgroups, we applied stratified sampling to maintain consistent distributions of race/ethnicity, PND prevalence across neighborhood areas where patients resided in each fold (**Fig.S2**). After data preprocessing, our final analysis included 62% Black, 8% White, 26% Hispanic, with an overall PND prevalence of 27%. See previous study for more details^22^.

### Identification of the important features

To identify the important features and understand the association with the severity of depressive symptoms, we applied the Shapley Additive Explanations (SHAP^28^ () and calculated the Shapley values. These values provide a quantification of the contribution of each feature in each patient to the model prediction. A higher absolute SHAP value indicates the feature had a greater influence on the model’s prediction performance. We also calculated cumulative Shapley values by summing the absolute SHAP values for features within each category (EMR biological, EMR sociodemographic, and neighborhood-level factors) to assess their relative overall importance to the model. We used Spearman correlation to estimate the direction of the effect between features (e.g., #EMR or #CHA) and PND outcome. See previous study for more details^22^.

### Bias assessment

We employed common metrics to assess bias: disparate impact (DI, eq.1)^29^, the equal opportunity difference (EOD, eq.2)^30^, the standard deviation of true positive and false positives (Equalized Odds [EO], eq.3,4) ^31^ across race/ethnicity.

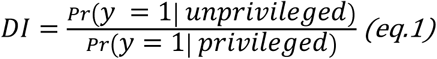

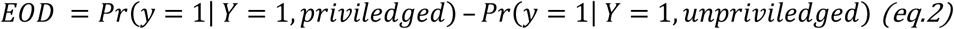

Here, *Y* and *y* denote the true and predicted outcome of depression (case=1, control=0), respectively. We selected NHW women as the privileged group, and NHB and Latina women as the unprivileged groups.

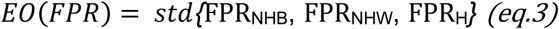

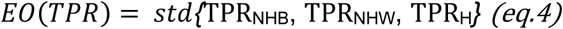

To further evaluate the contribution of each neighborhood level factor to the model bias, we employed a feature masking approach. In this method, each neighborhood level factor *i* (e.g., mortality rate) was masked one at a time by replacing it with its median value, while keeping the rest of the features unchanged. The pre-trained elastic-net model was then used to generate predictions on the masked dataset, and the bias metrics were calculated. The change in bias, denoted as ⍙*Bias_i_*, for each feature, *i* was assessed by comparing the fairness metrics, from equation 1 to 4, before and after masking, allowing us to identify features that contribute to each of the four bias metrics (eq.5).

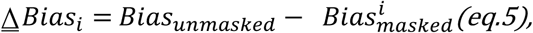

which *Bias* could be denoted as *DI, EOD, EO(TPR) or EO(FPR)*.

### Statistical analysis

The Mann-Whitney test was employed to assess ROCAUC and PRAUC differences across the three race/ethnic groups and the Wilcoxon signed-rank test to evaluate bias differences between models: the model using only EMR features, EMR with race/ethnicity included, and EMR with neighborhood level information without race/ethnicity; To determine the association of each feature with the outcome, Spearman correlation was used to estimate the direction and magnitude of the effect. All multiple comparisons were adjusted using the Bonferroni correction method.

### Geospatial analysis

Spatial representations of social neighborhood information and EMR were generated with the *geopandas* Python package (version 0.14.3), using the Chicago city map obtained from the Chicago Data Portal^32^. For representation purposes, we computed the rates (percentages) within the sample population of that area for categorical features, whereas continuous features were derived from computed mean values.

### Network analysis

Correlation networks were constructed to visualize relationships between CHA) features, EMR) variables, and PND risk. We calculated Spearman correlations among all variables using data from 3,313 individuals as well as within each racial/ethnic group separately. EMR variables were included in the network if their correlations with neighborhood-level variables were statistically significant (adjusted p < 0.05) after False Discovery Rate correction for multiple comparisons. Networks were created with *Cytoscape*^33^(version 3.10.2).

## RESULTS

### Incorporating social neighborhood information with EMRs while predicting PND improves model fairness

We extracted electronic health records and neighborhood-level data from a total of 6,137 pregnant patients who received obstetric care at a public urban hospital, with a total of 694 EMR and 260 neighborhood features. After data preprocessing, a total sample of 3,313 patients, 57 EMR and 125 neighborhood-level features were employed for downstream analysis.

Our predictive model for PND incorporated both neighborhood-level factors and EMR features and demonstrated moderate predictive performance across different racial/ethnic groups (ROCAUC: NHB (55%), NHW (57%), H (58%); PRAUC: NHB (32%), NHW (19%), H (26%), **Fig.2A,B**). Notably, the inclusion of neighborhood-level factors led to a significant reduction in bias, as evidenced by a decrease in the disparate impact (DI) and equalized odds false positive rate (EO(FPR)) (**Fig.2C,D,** p-value < 0.05) compared to models that just include EMRs or EMRs with race/ethnicity features. Furthermore, stratified sampling based on patients’ neighborhood residence effectively reduced performance variations across different neighborhood areas in NHW populations (**Fig.S3**).

**Figure 2.**
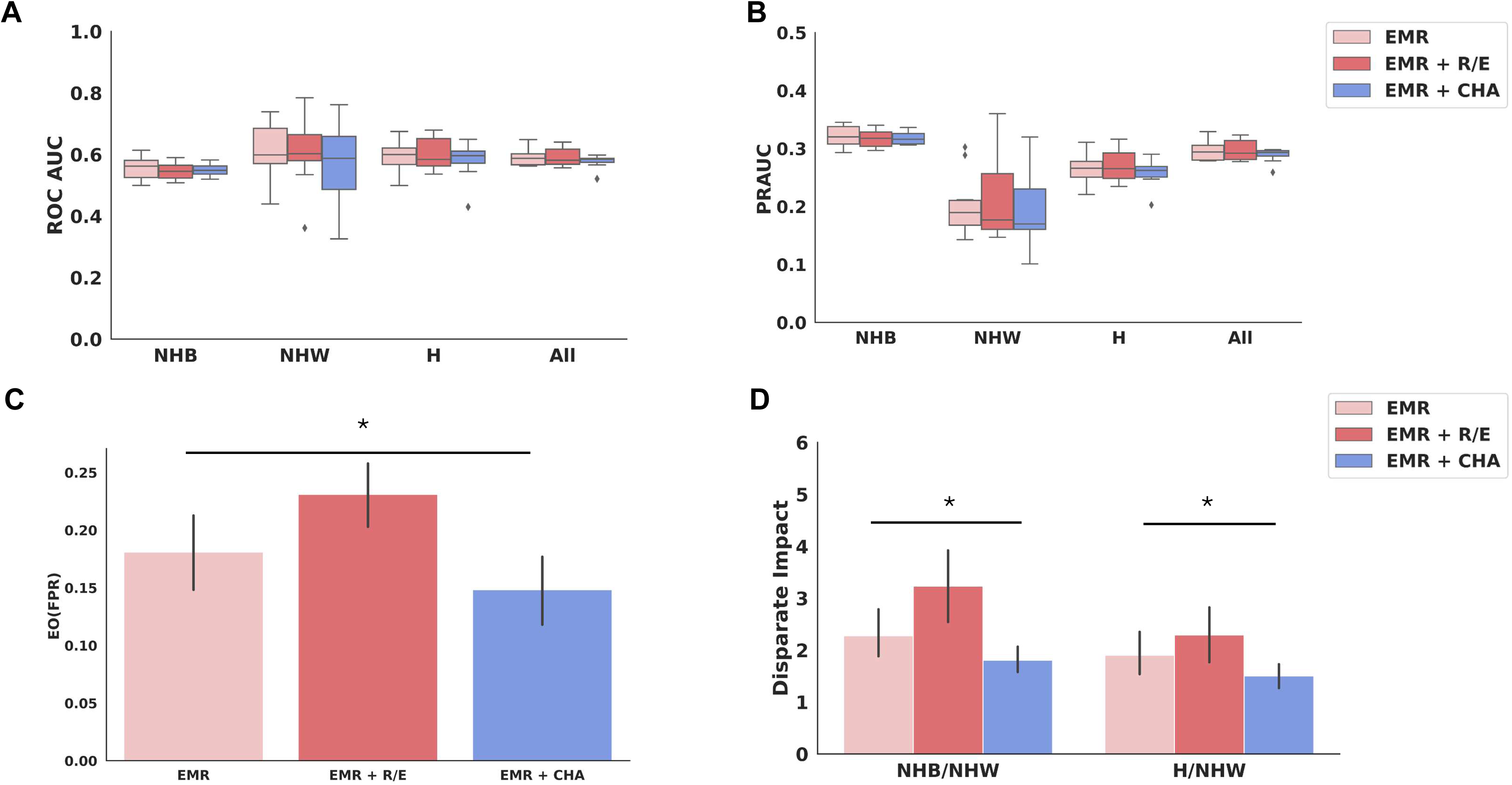
Model performance and fairness metrics across different racial/ethnic groups. Integration of Neighborhood-Level Data with Electronic Medical Records Enhances Machine Learning Model Fairness in Predicting Perinatal Depression While Maintaining Moderate Predictive Performance. **A,** ROC-AUC scores for Non-Hispanic Black (NHB), Non-Hispanic White (NHW), Hispanic (H), and overall population. **B,** Precision-Recall AUC scores across racial/ethnic groups. **C,** Equalized odds (false positive rate) comparison between models using EMR alone, EMR with race/ethnicity (R/E), and EMR with CHA data. (D) Disparate impact ratios comparing NHB/NHW and H/NHW across different model configurations. *p < 0.05.

### Identification of neighborhood-level factors for prenatal depressive symptom severity

To identify the features associated with depressive symptom severity early in pregnancy and the directionality of their associations, we calculated the Shapley values for each feature. Grounded in game theory, Shapley values provide an estimate of the unique contribution made by each feature towards the overall predictive performance of the model^28^. The top 25 EMR features with the highest mean absolute Shapley values included well-established sociodemographic risk factors for PND, such as having an unplanned pregnancy^34^; being single^35^, young^36^; covered by federal aid insurance^37^; having religion; use of alcohol, substance and tobacco^38^ (**Fig.3)**. These findings align with our previous study^22^, which unplanned pregnancy; being single; and covered by federal aid insurance as significant predictors of PND risk. The model also captures several health markers. For instance, elevated depressive symptomatology positively associated with self-reported levels of pain as well as pulse rate and the use of various medications, including antihistamines, anti-inflammatory/analgesics, antibiotics, and mood/anxiety medications. Additionally, common blood panel measurements, such as elevated white blood cell count (WBC), lower platelet levels (PLT), higher mean platelet volume (MPV), higher mean corpuscular hemoglobin (MCHC), and higher red cell distribution width (RDW) were identified as significant predictors (**Fig.3A**).

**Figure 3.**
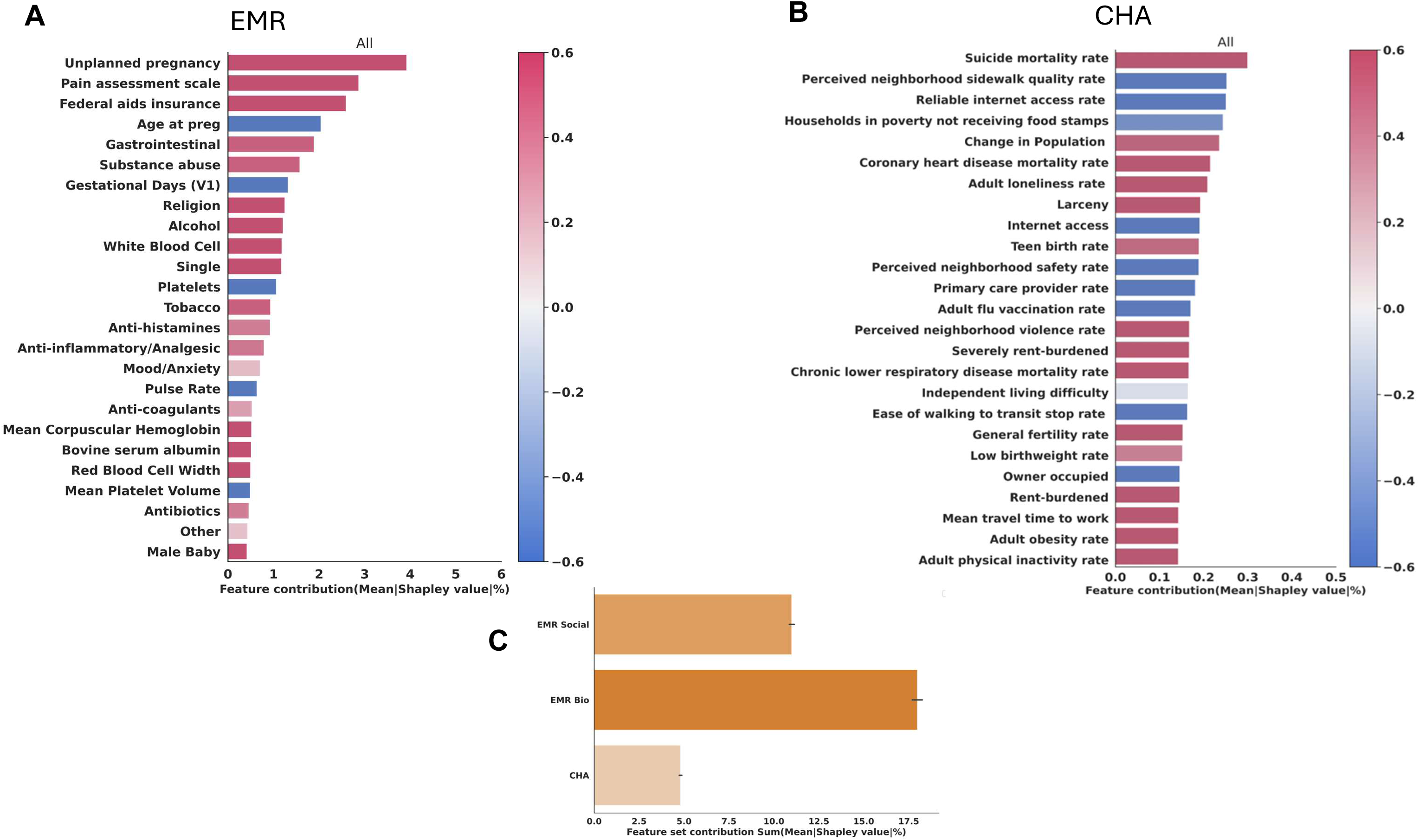
Electronic Medical Records better predicted the PND risk than most of the social neighborhood factors. **A,** Top EMR features ranked by mean SHAP values showing individual-level predictors of PND risk. **B,** Top neighborhood-level features ranked by mean SHAP values showing neighborhood-level predictors. **C,** Comparison of overall feature set contributions between EMR social factors, EMR biological factors, and neighborhood-level features. Color scale indicates positive (red) or negative (blue) association with PND risk.

The top 25 neighborhood level factors with the highest mean absolute Shapley values revealed significant neighborhood-level factors influencing PND risk (**Fig.3B**). Notably, the neighborhood suicide mortality rate has the highest Shapley value, followed closely by the perceived sidewalk quality rate. Other important neighborhood characteristics included reliable internet access, and poverty levels (particularly households not receiving food stamps despite meeting the poverty threshold). Population change rate in the neighborhood area over time, also played a significant role. Health-related factors such as coronary heart disease mortality rate, and adult flu vaccination rate were among the top contributors. Interestingly, adult loneliness rate, perceived neighborhood safety and violence rates, as well as factors related to housing (e.g., severely rent-burdened households) and accessibility (e.g., ease of walking to transit stops) also emerged as important neighborhood level predictors of PND risk. Overall, EMR biological features showed the highest cumulative Shapley values, followed by EMR sociodemographic features, while neighborhood level factors had a comparatively lower total contribution to the model predictions (**Fig.3C**).

### Neighborhood-level factors and their differential impact on PND risk across Race/Ethnicity

As neighborhood factors can modify biological characteristics and we observed that the major predictors of PND risk were EMR-associated features, we explored the association between the most predictive neighborhood-level factors and the most predictive EMR features of the PND risk. Our results revealed significant correlations encompassing both social demographics and biological factors (**Table S2-5**), with varying patterns of association across different racial and ethnic groups. NHB women displayed the highest number of significant correlations between EMR-based PND risk factors and neighborhood factors, followed by NHW individuals. And Hispanic women show fewer, but still notable, correlations (**Fig.4A-C**).

**Figure 4.**
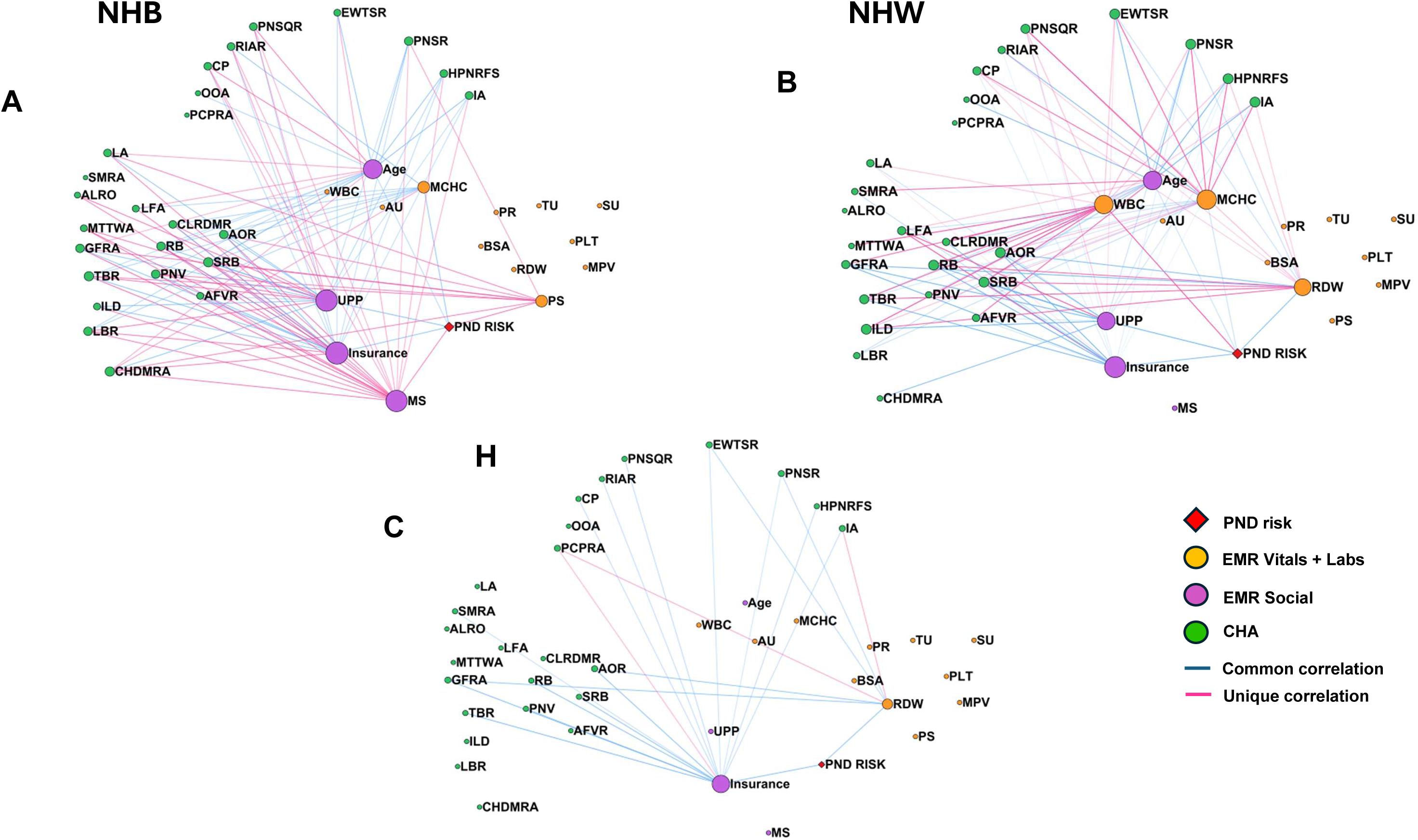
Network analysis of correlations between EMR and neighborhood-level factors across racial/ethnic groups. Network diagrams for **A,** Non-Hispanic Black **B,** Non-Hispanic White, and **C,** Hispanic populations showing relationships between PND risk (red diamond), EMR vitals/labs (orange circles), EMR social factors (purple circles), and neighborhood-level features (green circles). Blue lines indicate common correlations across groups, while pink lines show correlations unique to each racial/ethnic group. Node annotation was described in **Table S2**. Size of node indicates number of connections.

Neighborhood factors with strong correlations include indicators of resource scarcity (e.g., low internet access rates, limited food access, ease of walking to transit stops), access to healthcare measures (e.g., primary care provider rates), and health-related factors (e.g., adult obesity rates, suicide mortality rates). Socioeconomic indicators, particularly severe rent burden, display strong associations across groups, underscoring the significance of economic factors in health outcomes. For NHB individuals, negative social neighborhood factors have a more associations with social demographic features. Individual-level social demographic factors like unplanned pregnancy, being single, and young age were associated with several neighborhood-level characteristics such as higher mortality and morbidity rates, housing instability, poverty, higher levels of perceived violence, lack of accessibility to care, and resource scarcity. Biological health factors, such as higher pain assessment scores, were uniquely associated with higher levels of perceived violence. Lower MCHC correlated with higher mortality rates and low neighborhood rent burden (**Fig.4A**). For NHW individuals, neighborhood level factors had more influence on their biological health factors than in NHB and H, particularly higher levels of WBC, RDW and lower level of MCHC correlated with low neighborhood socioeconomic status and living difficulties. Similar to NHB, individual-level social demographic factors (unplanned pregnancy, being single, lower age) in NHW were associated with higher mortality or morbidity rates and low socioeconomic status (**Fig.4B**). Hispanic women showed fewer correlations overall, with connections primarily between use of federal aided insurance and neighborhood-level factors, such as rental burden, violence rate (**Fig.4C**). These findings highlight the complex interplay between individual-level risk factors and neighborhood-level social determinants of health, with notable variations across racial/ethnic groups.

### Quantifying bias contribution of neighborhood-Level factors in PND risk models across racial/ethnicity

Further analysis revealed differential impacts of neighborhood-level factors on model fairness. The effect of individual neighborhood features on the model biases showed that the addition of features such as suicide mortality rate led to the largest decreases in EO(FPR) difference, while adult obesity rate, severely rent-burdened households, and adult loneliness rate resulted in increases model biases (**Fig.5A**). Unexpectedly, when comparing the DI values using NHW as a reference, we observed that most neighborhood factors produced more bias toward NHB group (lower predictive capability) while reducing bias in H group (**Fig.5B**).

**Figure 5.**
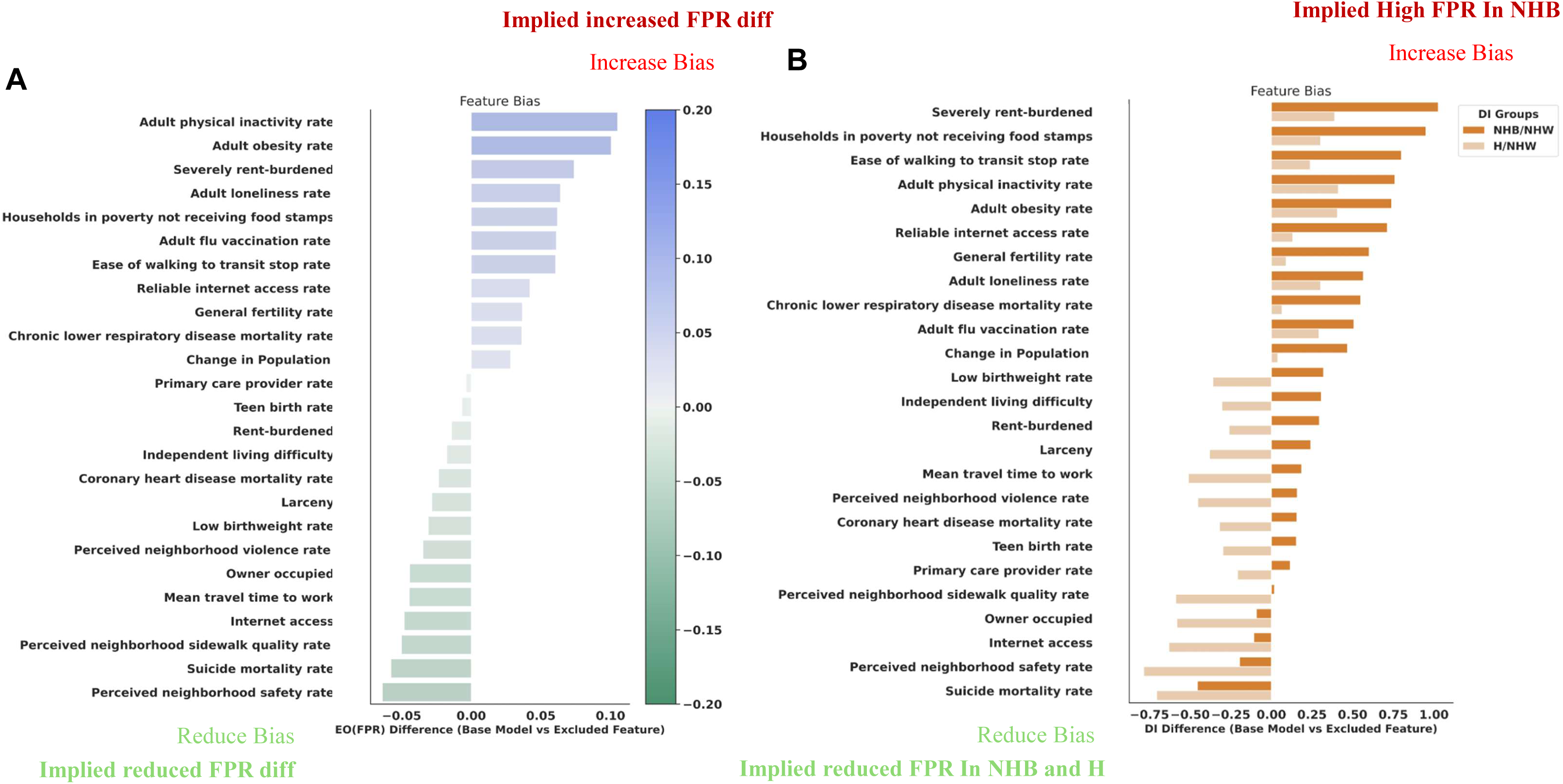
Impact of neighborhood-level factors on model bias. **A,** shows changes in equalized odds (EO) false positive rate (FPR) differences when individual neighborhood-level features are excluded from the model. **B,** shows changes in disparate impact (DI) ratios for NHB/NHW and H/NHW comparisons. Positive values indicate increased bias, while negative values indicate reduced bias. Features are ordered by their impact on bias metrics.

Several key neighborhood-level factors significantly reduced bias in prenatal depression prediction across racial/ethnic groups. The preceded neighborhood safety rate and suicide mortality rate demonstrated the strongest bias-reducing effects overall, while internet access and the owner-occupied housing rate consistently improved fairness across groups. Some factors showed group-specific benefits: primary care provider rate, coronary heart disease mortality rate, low birthweight rate, and the teen birth rate reduced bias particularly in Hispanic women, while perceived neighborhood safety improved fairness more generally. However, not all neighborhood factors were beneficial for fairness. Socioeconomic indicators, such as severely rent-burdened households and households in poverty not receiving food stamps, increased bias, especially for Black women. Similarly, certain health-related factors (adult obesity rate, adult loneliness rate) and demographic indicators (general fertility rate, population change) worsened disparities across groups. These findings highlight that while incorporating social neighborhood information can mitigate model bias in prenatal depression prediction, careful selection of specific neighborhood variables is essential, as their impacts vary substantially across racial/ethnic groups.

## DISCUSSION

We evaluated the ability of the addition of neighborhood-level measures to EMRs to improve the prediction of PND in early pregnancy of low-income women. In addition, we also investigated the bias contribution of neighborhood-level factors across racial and ethnic groups. Our results showed that the addition of neighborhood-level factors does not affect model performance but reduces bias. Among the top variables identified in our study were several neighborhood-level factors not previously associated with PND, including sidewalk quality, change in population, and neighborhood internet access rate. We also found associations with previously documented factors such as suicide mortality rate^39^, access to care^40^, food insecurity^41^, rental burden^42^, adult obesity^43^, teen birth and fertility rate, neighborhood violence, sidewalk quality, and transportation^44^. Taken together, the integration of neighborhood-level information increases the fairness of ML models while maintaining moderate PND prediction performance.

Our examination of associations between EMRs and neighborhood-level data reveals complex patterns in how neighborhood factors interact with EMR-related PND risk indicators and varied across racial and ethnic groups. NHB women displayed the highest number of significant correlations between PND risk factors and neighborhood-level factors, indicating the neighborhood-level information might more strongly mediate the relative risk of PND in NHB compared with NHW pregnant women. This may be due to the cumulative effects of chronic stress from multiple structural inequities and socioeconomic disparities, which can lead to allostatic load (the physiological “wear and tear” on the body from chronic stress exposure) and increased vulnerability to mental health disorders^45, 46^. NHW populations demonstrate links between both individual social demographics and biological factors (such as higher WBC, RDW and lower MCHC) with neighborhood-level factors, indicating that environmental stressors may have both psychosocial and physiological impacts on PND risk in this group.

Despite facing similar adverse neighborhood conditions, Hispanic individuals might be sheltered from the negative impacts of their neighborhood characteristics, given fewer correlations between neighborhood factors and PND risk indicators compared to other groups. This may imply that Hispanic women benefit from protective cultural factors such as familism (a cultural value emphasizing strong family bonds and support), and social support networks, potentially buffering against neighborhood-level stressors^47, 48^. However, this could also suggest underdiagnosis or underreporting of symptoms due to cultural views about mental health disclosure and help-seeking behaviors or language barriers^49^. In summary, our findings highlight the significant variation in risk factors and social determinants of health across racial/ethnic groups, thus the importance of including in the models to diagnose PND.

Neighborhood-level factors show varied impacts on ML model bias across racial groups, as measured by the Equalized Odds False Positive Rate and Disparate Impact ratio. Neighborhood-level features such as “severely rent-burdened households”, and “households in poverty not receiving food stamps” showed the highest increase in model bias across both metrics when included in the ML models. This pattern likely reflects the systemic inequalities in resource allocation and economic opportunities that disproportionately affect minority communities,^50^ suggesting that economic stress may be particularly affecting how our model identifies depression risk during pregnancy for these communities. Neighborhood-level health-related factors, including adult obesity, adult loneliness, general fertility and chronic lower respiratory disease mortality rates, also contributed to increased bias, possibly due to the compounded effects of limited healthcare access and environmental stressors in disadvantaged neighborhoods^46^. Interestingly, neighborhood safety and the suicide mortality rate demonstrated the strongest bias-reducing effects, as evidenced by their negative EO(FPR) differences and reduced DI ratios. This finding might indicate that these factors could be more universal indicators of neighborhood distress that affect all racial groups more uniformly, thereby leading to more balanced risk assessments^9^. Furthermore, most of the neighborhood-level features increased bias particularly in (NHB) groups compared to Hispanic (groups, which may denote the cumulative effects of structural racism and economic inequality on health outcomes in NHB communities^50^. In summary, these findings highlight how neighborhood-level factors might impact the detection of PND symptoms differently across racial and ethnic groups, emphasizing the need for culturally sensitive approaches in both PND screening and prediction models.

Our study demonstrates several novel contributions to the field of prenatal depression prediction and healthcare equity. Firstly, we successfully enhanced model fairness while maintaining predictive accuracy by incorporating neighborhood-level information into our machine learning models. This approach addresses a critical gap in current PND prediction methods, which often overlook the broader social context of patients, particularly those from disadvantaged neighborhoods. Secondly, our network analysis provides a unique perspective on the influence of neighborhood-level factors on patient health, specifically in relation to PND risk. This comprehensive view reveals complex interconnections between social determinants of health and individual risk factors, offering a more nuanced understanding of PND risk across different racial and ethnic groups. Lastly, identifying specific neighborhood-level factors that either increase or reduce model bias represents a significant step towards developing more equitable healthcare prediction tools.

Our study has several limitations that should be addressed in future research. Firstly, the sample size of NHW was relatively small, which may affect the generalizability of our findings and the statistical power of our analyses. Secondly, while our integration of neighborhood-level information with EMRs represents an important step forward, more advanced integration methods should be developed to fully leverage the potential of combining these data sources. Additionally, our study was geographically limited, which may restrict the applicability of our findings to other regions with different social and environmental contexts. Future research should aim to validate these findings in diverse geographic and demographic contexts and explore interventions that leverage neighborhood-level factors to reduce disparities in PND risk and care.

## CONCLUSION

Our study demonstrates that incorporating neighborhood-level information into machine learning models for prenatal depression prediction not only enhances model fairness but also maintains predictive accuracy. Through comprehensive analysis of neighborhood-level factors, we identified key features that reduce as well as increase model bias. The differential impact of these factors across racial and ethnic groups highlights the complex interplay between neighborhood conditions and individual risk factors. These findings emphasize the importance of considering neighborhood context in clinical risk assessment and suggest that addressing neighborhood-level factors could be crucial for reducing disparities in prenatal depression care. Future research should explore additional data sources within the Chicago area to validate these findings and explore interventions that leverage neighborhood-level factors to improve maternal mental health outcomes.

**Table 1.** Demographic and clinical characteristics of the study population stratified by PND status and race/ethnicity. The study included 3,313 pregnant women (843 cases, 2,470 controls) from a public hospital after preprocessing. NHB: non-Hispanic Black. NHW: non-Hispanic White. H: Hispanic or Latina.

| N, Percentage |  |  |  |  |  |  |  |  |  |  |
| --- | --- | --- | --- | --- | --- | --- | --- | --- | --- | --- |
|  | Case<br>(N = 843) |  |  |  |  | Control<br>(N = 2470) |  |  |  |  |
|  | All cases | NHB | NHW | H | Other | All controls | NHB | NHW | H | Other |
|  |  | 594(70%) | 39(5%) | 192(23%) | 18(%) |  | 1465(59%) | 222(9%) | 666(27%) | 117(5%) |
| **Single** | 716(85%) | 543(91%) | 18(46%) | 151(79%) | 4(22%) | 1846(75%) | 1285(90%) | 86(39%) | 439(66%) | 36(31%) |
| **Federal Aided Insurance** | 698(83%) | 512(86%) | 19(49%) | 161(84%) | 6(33%) | 1742(71%) | 1160(79%) | 68(31%) | 475(71%) | 39(33%) |
| **Unplanned pregnancy** | 679(81%) | 515(87%) | 23(59%) | 138(84%) | 3(17%) | 1566(63%) | 1125(77%) | 53(24%) | 358(54%) | 30(36%) |
| Median, Min ,Max |  |  |  |  |  |  |  |  |  |  |
| **Age** | 26(13-47) | 25(13-43) | 30(16-43) | 26(15-47) | 29(24-41) | 27(13-50) | 26(13-50) | 31(18-45) | 26(14-46) | 30(17-44) |
| **BMI** | 28(15-212) | 28(16-81) | 25(19-50) | 29(15-212) | 24(19-35) | 28(11-155) | 29(17-155) | 25(17-83) | 29(11-57) | 24(16-46) |
| **PHQ-9** | 12(10-27) | 11(9-27) | 11(9-24) | 11(9-27) | 12.5(10-19) | 2(1-4) | 2(1-4) | 2(1-4) | 2(1-4) | 2(1-4) |

## Data Availability

Data is available to the research community upon approval of the University of Illinois Chicago Institutional Review Board (IRB # 2020-0553).

## Acknowledgments

This work is funded through NICHD (R21HD110779). BPB has been supported by NIMH (U54MD012523). Additionally, this work has been also supported by the National Center for Advancing Translational Sciences (NCATS), National Institutes of Health, through Grant Award Number UL1TR002003. We would like to thank Dr. Subhash Kumar Kolar Rajanna for his assistance in extracting the electronic medical records used in this manuscript. The content is solely the responsibility of the authors and does not necessarily represent the official views of the NIH.

## Author contributions

BPB and YD conceptualized the idea; BPB, YD, and YH designed the methodology for model creation and validation; BPB, YH, and SA curated and pre-processed the data; YH performed the analysis and subsequent validation; BPB, YD, YH, SJK and PMM interpreted the results and wrote the original draft; and all the authors critically reviewed and edited the manuscript.

## SUPPLEMENT FIGURE AND TABLE LEGEND

**Table S1. EMR data preprocessing.** Breakdown of number of patients through the preprocessing steps.

**Table S2. Annotation of variable in network.**

**Table S3.Significant correlation among EMR, neighborhood level factors and PND risk in NHB.** Significant correlations among individual EMR, neighborhood-level features, and PND risk in Non-Hispanic Black (NHB) participants. The “Edge name” column indicates the pair of variables being correlated, “Correlation Coef” shows the strength and direction of the correlation, and “Edge Type” indicates whether the correlation is unique or repeated across race/ethnicity.

**Table S4.Significant correlation among EMR, neighborhood level factors and PND risk in NHW.** Significant correlations among individual EMR, neighborhood-level features, and PND risk in Non-Hispanic Black (NHB) participants. The “Edge name” column indicates the pair of variables being correlated, “Correlation Coef” shows the strength and direction of the correlation, and “Edge Type” indicates whether the correlation is unique or repeated across race/ ethnicity.

**Table S5.Significant correlation among EMR,** neighborhood **level factors and PND risk in H.** Significant correlations among individual EMR, neighborhood-level features, and PND risk in Non-Hispanic Black (NHB) participants. The “Edge name” column indicates the pair of variables being correlated, “Correlation Coef” shows the strength and direction of the correlation, and “Edge Type” indicates whether the correlation is unique or repeated across race/ ethnicity.

**Supplement Figure 1. Nested k-fold cross validation with neighborhood level stratification.** Outer loop consists of 10 folds, which split the data into train(yellow) and test(green) set. Inner loop consists of 5 folds, which split train data from the outer loop into another train(yellow) and validation(green) set. Each fold were stratified based on race/ethnicity and case/control. NHB: non-Hispanic Black. NHW: non-Hispanic White. H: Hispanic or Latina.

**Supplement Figure 2. Disparity in Non-Hispanic White population**. **A,** Group 1: NHW ROCAUC > 61%, better predicted NHW cases are mainly from south and west side of the Chicago. **B,** Group 2: NHW ROCAUC < 61%, worst predicted NHW cases are mainly from north and north-west of Chicago. **C,** Overall patient distribution across Chicago neighborhoods. These variations likely emerged due to the relatively small NHW population in our cohort (9% of total, **Table 1**) and their uneven distribution across Chicago neighborhoods, which could lead to unstable model performance for this group.

**Supplement Figure 3. Predictive performance assessment of models before and after applying stratified sampling.** Model performance before (**A, B**) and after (**C, D**) applying stratified sampling to ensure consistent distributions of race/ethnicity and PND prevalence across neighborhood areas in each fold. (**A, C**) ROCAUC curves and (**B, D**) PRAUC curves for each racial/ethnic group and overall (All). ns: not significant; *, **: significant differences (p < 0.05, p < 0.01). **EMR:** Model with EMR alone; **EMR + R/E**: EMR and race/ethnicity; **EMR + CHA**: EMR and neighborhood level factors; **EMR + top CHA**: EMR and top neighborhood level factors.

