## supplement figures for "Leveraging neighborhood-level Information to Improve Model Fairness in Predicting Prenatal Depression"

### Slide 1
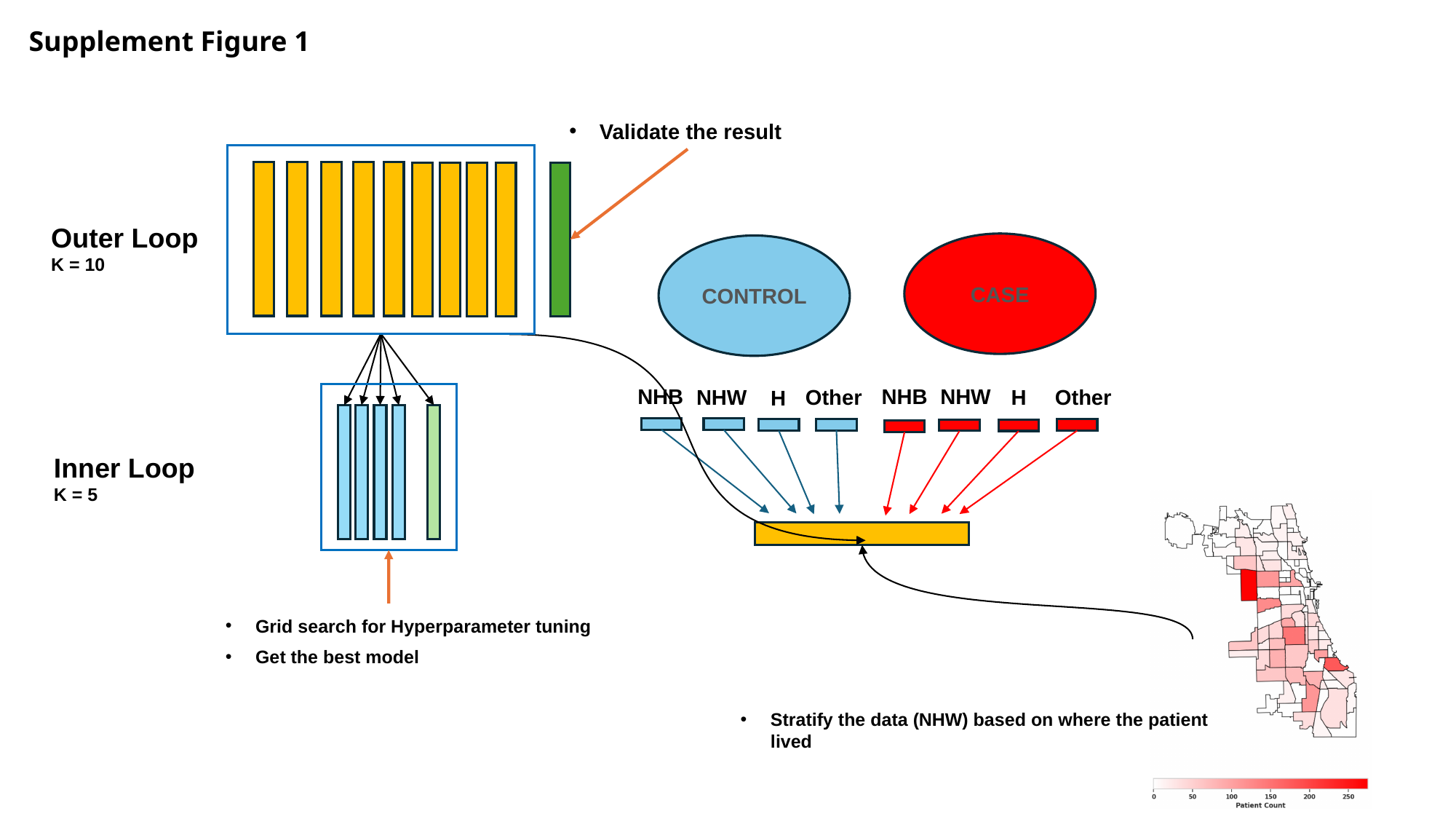

Supplement Figure 1
Validate the result
Outer Loop
K = 10
CASE
CONTROL
NHB
NHB
NHW
Other
H
Other
NHW
H
Inner Loop
K = 5
Grid search for Hyperparameter tuning
Get the best model
Stratify the data (NHW) based on where the patient lived

### Slide 2
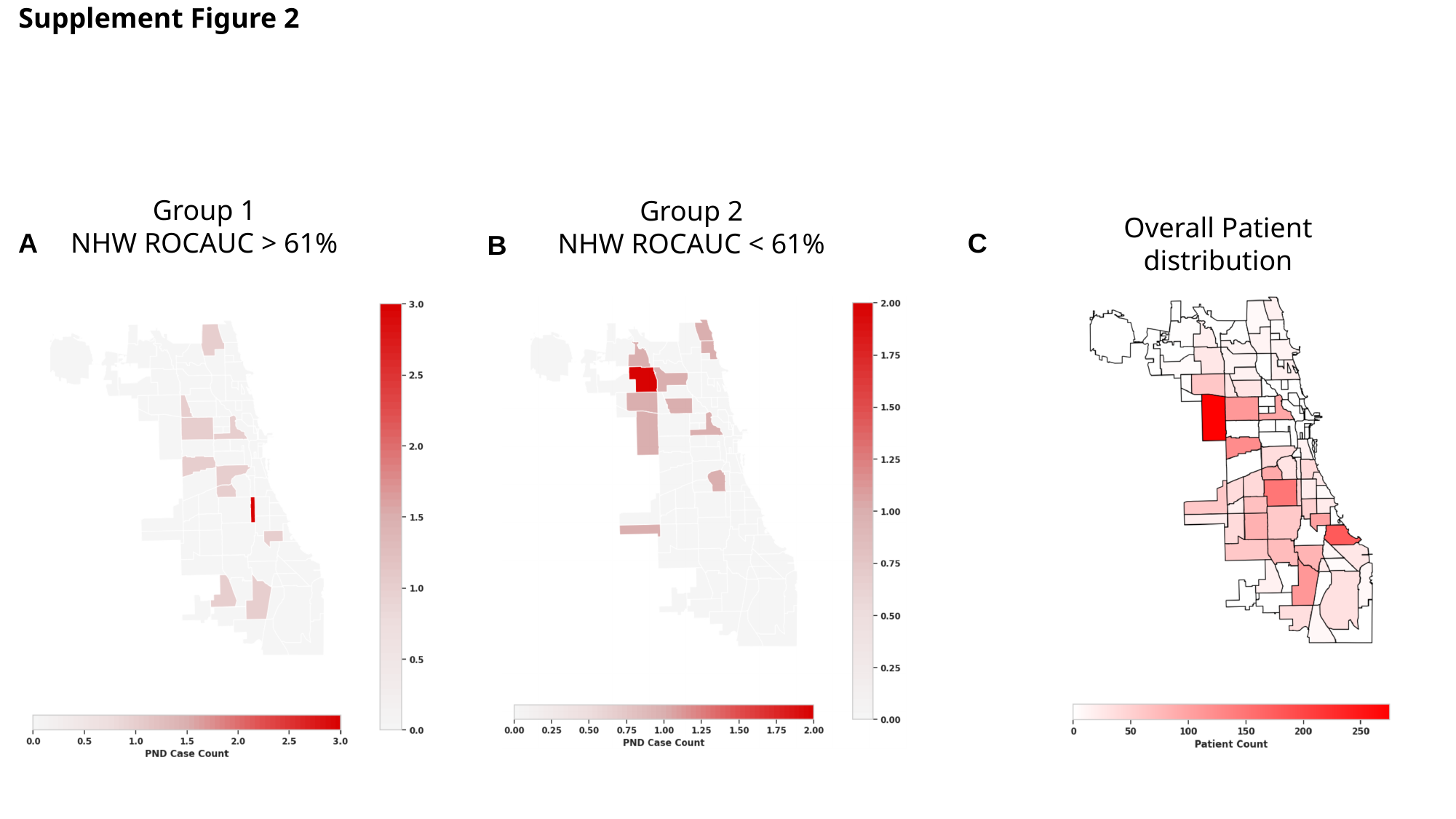

Supplement Figure 2
Group 1
NHW ROCAUC > 61%
Group 2
NHW ROCAUC < 61%
Overall Patient distribution
A
C
B

### Slide 3
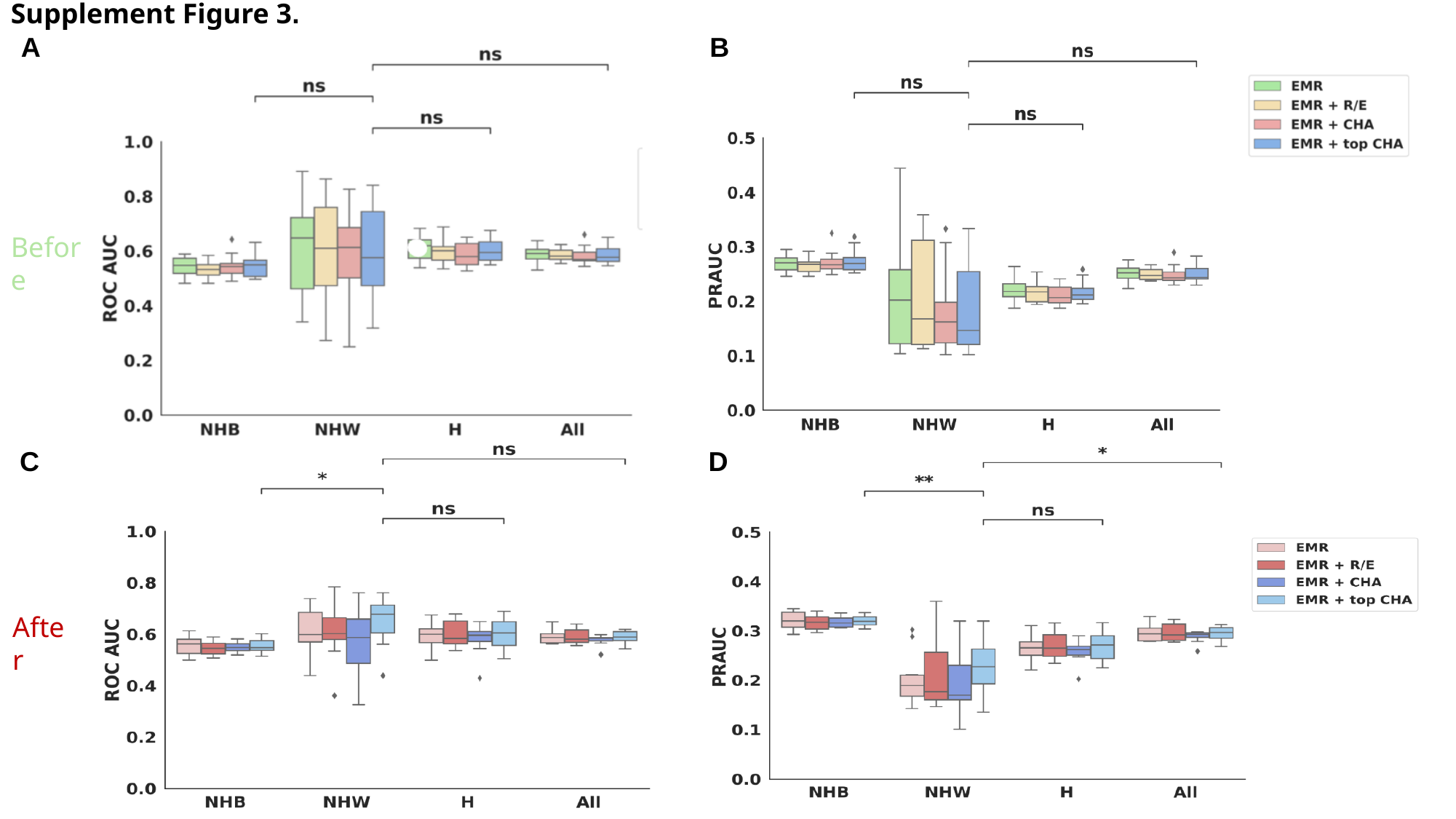

Supplement Figure 3.
A
B
Before
C
D
After
